# Neural Correlates of Reward Dysfunction in Adolescent Cannabis Use and Depression

**DOI:** 10.1101/2025.11.01.25339301

**Authors:** Tram N. B. Nguyen, Benjamin A. Ely, Emily R. Stern, Russell H. Tobe, Vilma Gabbay

## Abstract

**Objective:** Cannabis use is prevalent among youth with depression and may alter the reward neurocircuitry, which also plays a role in depression. Yet, reward function in co-occurring cannabis use and depression remains poorly understood. Here, we sought to examine neural reward processing in an adolescent sample with varying cannabis use and depression severity.

**Methods:** Participants completed diagnostic interviews and self-reported depression symptoms dimensionally. Cannabis use patterns were determined from clinician interviews, self-reports, and urine toxicology screens. Structural and Reward Flanker Task functional MRI data were collected and preprocessed using Human Connectome Project-style pipelines. Using a parcellated, network-based approach, we examined neural responses during reward expectancy (reward vs. non-reward cues) and attainment (reward vs. non-reward feedback) in relation to cannabis use and depression severity, controlling for age, sex, and multiple comparisons.

**Results:** In the full sample of 117 adolescents (age: 15.5 ± 2.3 years, 63.3% female), greater depression severity was linked to blunted caudate activity during reward expectancy and heightened activation of posterior cingulate and entorhinal cortices during reward attainment. Among 34 adolescents who used cannabis, heavier use was associated with greater activation in the left mediodorsal posterior thalamus and habenula during reward expectancy, while depression severity positively correlated with activation across cortico-striatal, default, memory, and visual networks during reward attainment. Cannabis use × depression interaction effects were detected in frontal and entorhinal cortices during reward attainment. Exploratory analyses showed sex differences.

**Conclusions:** Our results reveal divergent neural profiles of cannabis use and depression, as well as their additive associations with altered reward processing in adolescents.

## 1. Introduction

Cannabis has surpassed alcohol as the most commonly used illicit substance among adolescents in the United States ^1^, coinciding with declining risk perception ^2^ amid policies relaxing regulations on cannabis products ^3^. These trends are concerning, as adolescence is a critical neurodevelopmental period in which reward circuitry is highly sensitive to disruption by exogenous cannabinoids ^4–6^. In rodents, juvenile exposure to Δ^9^-tetrahydrocannabinol (THC), the primary psychoactive compound in cannabis, can irreversibly perturb cortical-striatal reward pathways and induce depressive-like behaviors into adulthood ^7–9^. In humans, adolescent cannabis use increases the risk for future depression ^10–13^, and adolescents with depression are more than twice as likely to use cannabis ^14^ and progress to disordered use ^15,16^. Efforts to probe reward processes in adolescent cannabis use via functional magnetic resonance imaging (fMRI) have been limited, with existing studies showing modest evidence of neural alternations during reward expectancy and attainment ^17^. However, a recent meta-analysis of cannabis use in youth suggested that neural vulnerabilities in reward processing may be concentrated in high-risk groups (e.g., those with heavy cannabis use or comorbid psychopathology, including depression) ^18^. Meanwhile, fMRI studies on adolescent depression have consistently documented blunted striatal activity during both reward expectancy and attainment, while alterations in medial- and dorsolateral-prefrontal regions vary across cohorts ^19–21^. These findings implicate reward dysfunction as a shared neurobiological pathway linking adolescent cannabis use and depression. Relatedly, a concerning clinical phenomenon is the high prevalence of cannabis use among depressed adolescents ^14^, which may lead to worsening of reward dysfunction. There has been sparse research examining reward function in adolescents with depression and cannabis use.

Additionally, methodological considerations are important when examining adolescent reward function. Commonly used reward fMRI tasks predetermine outcomes and rely on simple choices ^22^, which may be less sensitive to developmentally salient motivational processes ^23^. We thus developed the reward flanker task (RFT), which combines elements of the Monetary Incentive Delay ^24^ and Flanker ^25^ tasks to enhance participant engagement. Notably, this effort-based task design allowed us to capture distinct neural responses to reward expectancy and attainment while incorporating cognitive control processes ^26,27^. In adolescents with diverse internalizing symptoms, we found that depression severity was associated with altered cingulate, insula, and striatal activation during reward attainment ^27^. We subsequently examined anhedonia, a clinical manifestation of reward dysfunction ^28,29^, in an expanded adolescent sample with varying cannabis use and depression severity. Adolescents with cannabis use endorsed greater anticipatory anhedonia, which reflects motivational processes related to reward expectancy ^29^, but not consummatory anhedonia compared to peers without cannabis use, despite comparable depression levels ^30^. These findings implicate deficits in motivation, rather than capacity to experience rewards, in adolescent cannabis use.

Building on this literature and our prior work, here we utilized the RFT to parse neural correlates of cannabis use and depression during reward expectancy and attainment. We hypothesized that: 1) Cannabis use would be associated with altered neural processing during reward expectancy; 2) Depression would be associated with altered neural processing during both reward expectancy and attainment; 3) There would be an interaction effect between cannabis use and depression on neural alterations across both reward phases. As substantial sex effects have been documented in both adolescent cannabis use ^18^ and depression ^31^, we also explored sex differences.

## 2. Methods

### 2.1. Recruitment and Initial Evaluations

We recruited participants in the New York metropolitan areas from October 2013 to August 2023. Participants and their families responded to our community advertisements or were referred to our research group by clinicians. The Institutional Review Boards (IRB) approved our study protocol at all research sites, including academic-affiliated health systems and a state-sponsored psychiatric research institute. Participants were first screened via brief phone interviews. Prior to any other study procedures, verbal assent was obtained from participants younger than 18 years, and informed consent was signed by their accompanying parent or guardian. Participants who were 18 years and older provided informed consent.

To guide our psychiatric assessments, we used the Kiddie Schedule for Affective Disorders and Schizophrenia – Present and Lifetime Version for Children (K-SADS-PL)^32^ for participants younger than 18 years and their accompanying parent as well as the Mini International Neuropsychiatric Interview (MINI) ^33^ for participants 18 years and above. Clinicians within our research group were trained on both instruments to determine the current and past psychiatric symptomatology and diagnoses of all participants. To rule out psychiatric presentation secondary to underlying medical conditions, we conducted blood tests assessing for complete blood count, thyroid function, and liver function. Additionally, on the neuroimaging scan day, participants underwent a urine toxicology test to screen for the presence of amphetamines, barbiturates, benzodiazepines, cocaine, 3,4-Methylenedioxymethamphetamine (MDMA), methamphetamine, opiates, oxycodone, propoxyphene, and THC. Information from the final clinical profiles as well as blood and urine tests were employed to determine study eligibility based on the criteria below.

### 2.2. Inclusion and Exclusion Criteria

All participants included in this study met the following criteria: a) age between 12 and 21 years; b) Tanner stage ≥4; c) post-menarche if female. Participants were excluded from this study if they had any of the following: a) a history of schizophrenia and other psychotic disorders, neurodevelopmental disorders, or any substance use disorder that was not CUD; b) current illicit use of stimulants, sedatives, opioids, or psychedelics, as reported on assessments or confirmed by urine toxicology screenings; c) an estimated full-scale IQ below 80, as assessed using the Kaufman Brief Intelligence Test (K-BIT) ^34^; d) medical conditions, including neurological disorders, as identified through medical history and blood tests; e) contraindications for MRI, f) a positive pregnancy test result. Individuals unable to reliably complete study procedures based on the judgment of the investigators were also excluded. Participants taking prescribed stimulant medication discontinued use a week before the scan. There were no exclusions related to sex, race, ethnicity, cannabis use, or depression severity. This dimensional approach enabled us to comprehensively assess the comorbidity of cannabis use and depression in adolescence without the constraints of categorical diagnoses ^35^.

### 2.3. Cannabis Use and Depression Assessments

Our clinical variables of interest were cannabis use and depression severity. To describe patterns of cannabis use, we incorporated data from clinician interviews, self-reports, and urine screens. As in prior work ^30^, cannabis use patterns were characterized into five levels: 0 = never used, 1 = tried once, 2 = low use, 3 = moderate use, and 4 = heavy use. Adolescents who had never used cannabis or had only tried it once were classified as adolescents without cannabis use. To capture depression dimensionally, we used the Children’s Depression Rating Scale-Revised (CDRS-R) ^36^, which contains 17 items and has demonstrated high reliability and validity in adolescents ^37^. When interviewing participants and their guardian separately using the CDRS-R, trained clinicians generated a clinical summary severity rating of each symptom specified by each item on a 1-to-5 or 1-to-7 scale, resulting in a possible total score range of 17 to 113. Higher scores on the CDRS-R denote more severe depression.

To further characterize participants’ clinical profiles (see **Table 1**), we measured relevant symptoms with the Multidimensional Anxiety Scale for Children (MASC) ^38^, Beck Scale for Suicide Ideation (BSSI) ^39^, and Temporal Experience of Pleasure Scale (TEPS)^40^. Anxiety, suicidality, and anhedonia levels were not further analyzed as they were not the focus of the present study. Additional details on cannabis use and symptom assessments are provided in the **Supplementary Methods**.

**Table 1.**
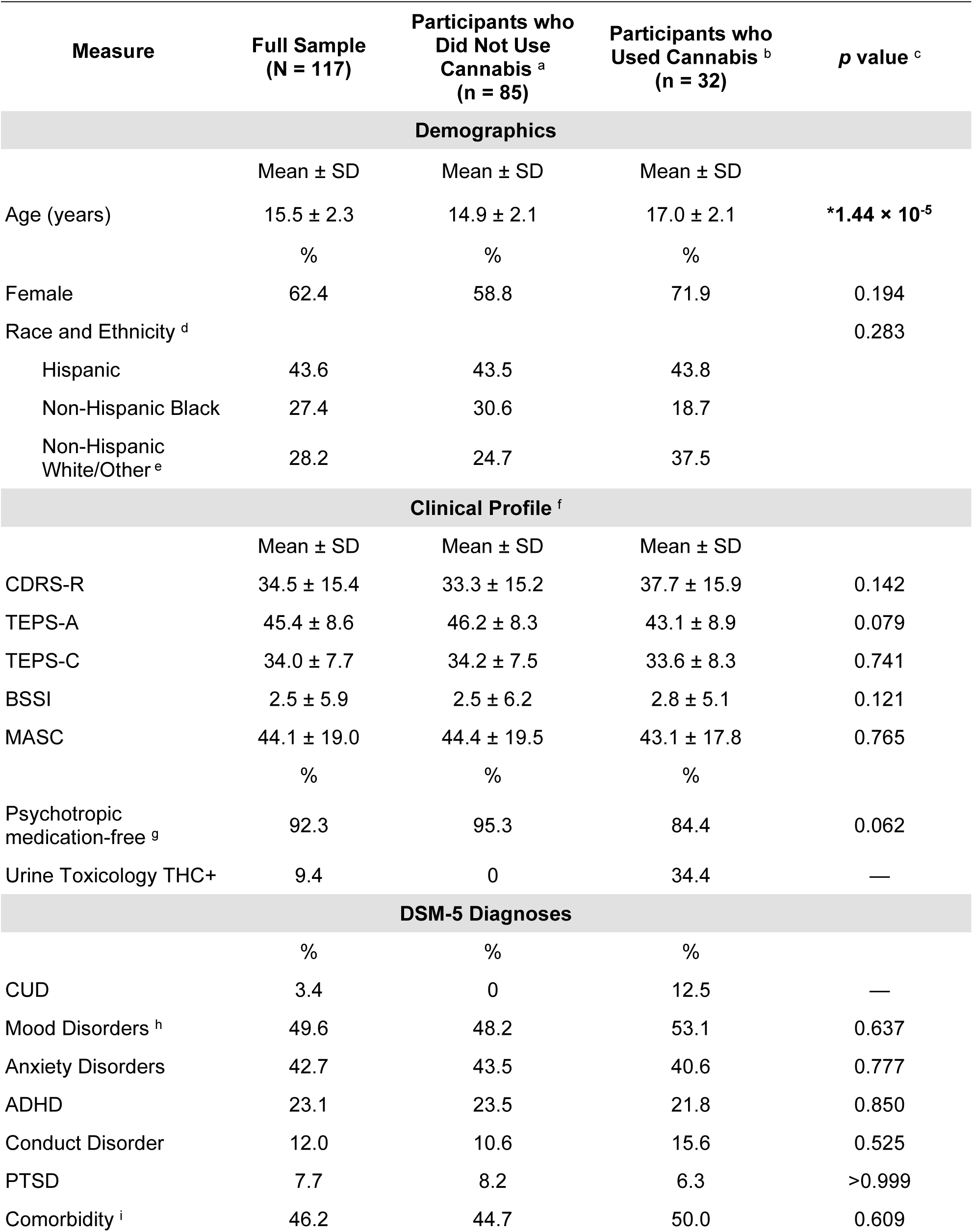
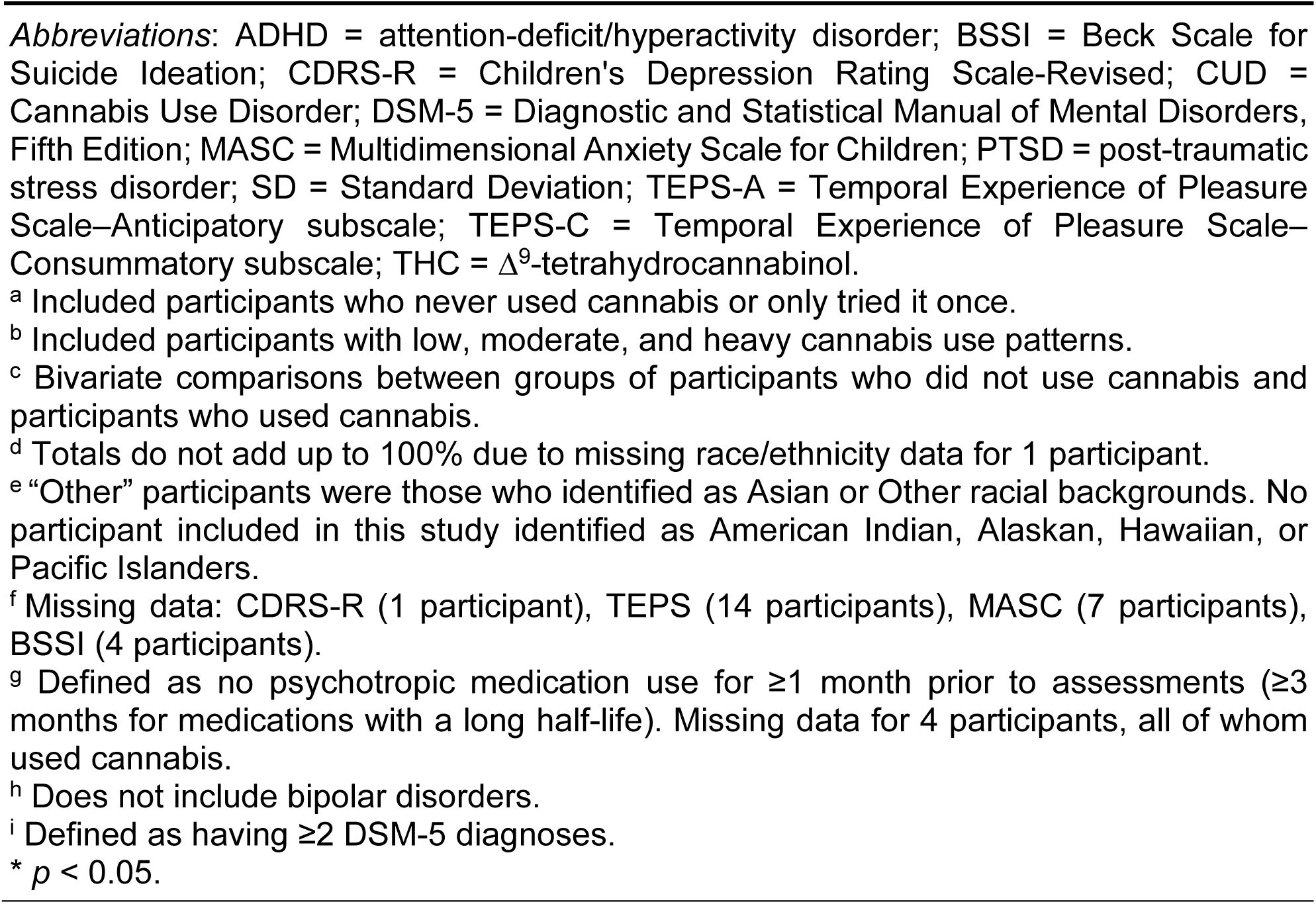
Demographic and clinical characteristics of study participants.

### 2.4. Magnetic Resonance Imaging (MRI) Acquisition

All participants completed a standard MRI safety screening prior to scans. Neuroimaging data were collected on a 3T Siemens Skyra scanner (Siemens, Germany) using acquisition protocols consistent with those implemented in the Human Connectome Project (HCP) Lifespan study ^41^. Most participants were scanned with a 16+4 channel head/neck receive coil, while 10 participants were imaged with a 32-channel head receive coil. Coil type was controlled for as a nuisance variable in all analyses.

Structural imaging consisted of a high-resolution T1-weighted MPRAGE sequence (TR = 2400 ms, TE = 2.06 ms, TI = 1000 ms, flip angle = 8°), acquired with 224 sagittal slices, no gap, a 256 × 256 matrix, a 230 × 230 mm^2^ field of view (FOV), and 0.9 mm isotropic voxels. In addition, a T2-weighted SPACE sequence (TR = 3200 ms, TE = 565 ms, flip angle = 120°) was collected, matched to the T1 acquisition in slice number, matrix size, FOV, and voxel resolution.

Functional MRI data were collected while participants performed 3-4 runs of the Reward Flanker Task (RFT; see below), each lasting 6 minutes and 14 seconds (374 volumes per run). Images were acquired with a T2*-weighted multiband echo-planar imaging (EPI) sequence at 2.3 mm isotropic resolution, using alternating left-to-right (LR) and right-to-left (RL) phase-encoding directions. Acquisition parameters included: TR = 1000 ms, TE = 31.4 ms, flip angle = 60°, 60 transverse slices (2.3 mm thickness, no gap), in-plane resolution = 2.3 × 2.3 mm, FOV = 624 × 720 mm, and multiband acceleration factor = 5. To support distortion correction, spin-echo EPI fieldmaps were also obtained in both LR and RL phase-encoding directions with matched acquisition parameters.

### 2.5. Reward Flanker Task

The event-related Reward Flanker Task (RFT) was described previously by our group ^26,27,42–44^ and others ^45,46^. Each trial presented a monetary cue (4-6 s), followed by a 300 ms flanker stimulus requiring a button press within a defined response window (≤1700 ms) to identify the central target. Response windows were individualized based on performance during a pre-scan training session, described below. Feedback (2 s) was then presented indicating trial accuracy (correct, incorrect, too slow) and monetary reward. Trials included certain (0¢, 10¢, 50¢) or uncertain (?) cues, with the latter linked to each reward level equally. On each trial, the interval between cue and feedback was varied to be between 4 and 6 s (plus reaction time) in order to break collinearity between cue and feedback events. Each RFT run included 30 pseudo-randomized trials. See **Figure 1** for example RFT trials.

**Figure 1.**
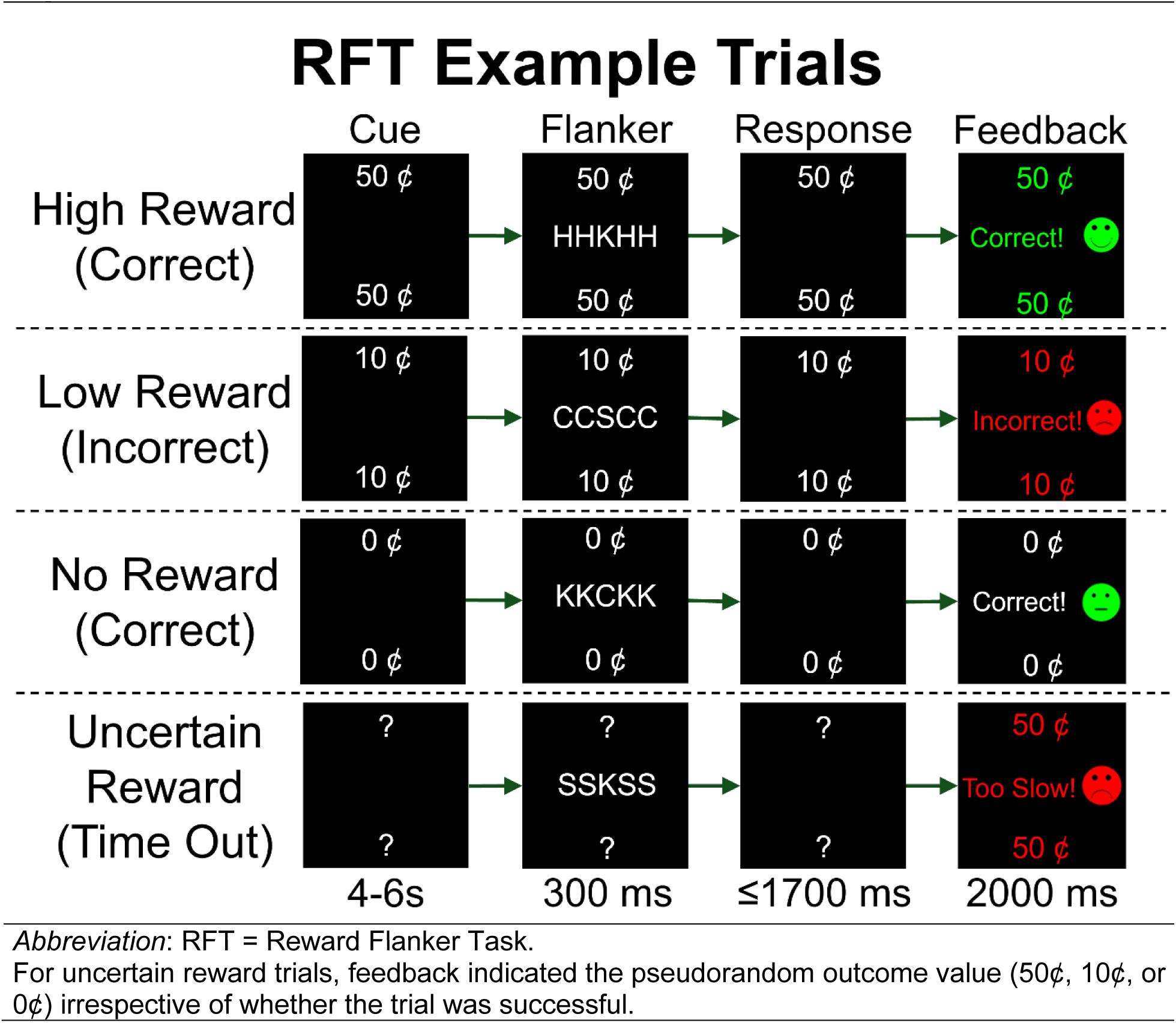
Example trials of the Reward Flanker Task.

On neuroimaging scan day, participants completed a brief training in a pediatric-friendly mock scanner to practice the RFT and acclimate to the scanning environment. This approach improves data quality, particularly in pediatric imaging ^47^. Mean response time (RT) from the mock session was used to calibrate the in-scanner task, with response windows scaled to 1.5 times the participant’s average practice RT, up to the task limit of 1700 ms.

### 2.6. MRI Data Preprocessing

All MRI data were visually inspected then processed with version 3.2 of the Human Connectome Project (HCP) Pipelines ^48^. This pipeline incorporated corrections for gradient nonlinearity, EPI distortions, AC-PC alignment, co-registration, and normalization to the Montreal Neurological Institute 152 non-linear 6th-generation (MNI152NLin6) space ^49^. Anatomical scans additionally underwent brain extraction, bias field correction, and cortical surface reconstruction with FreeSurfer, including cortical ribbon generation for grayordinate-based analyses. Functional data received similar corrections, motion realignment, spatial normalization, and intensity scaling before projection to 32k CIFTI grayordinate space, which preserves cortical and subcortical representation in a unified format ^48^.

Participants were excluded from further analyses if ≥2 RFT runs demonstrated excessive motion, characterized by more than 3% of frames exceeding 1 mm of relative displacement. To control for structured fMRI noise, we applied spatial independent component analysis (ICA) to whole-brain MNI-normalized timeseries, followed by automated component classification using the multi-run ICA-FIX framework ^50^. This approach leverages multiple resting-state and task runs to improve denoising for shorter scans ^51^. The FIX algorithm categorized components as “signal”, “noise”, or “unknown”. Trained neuroimagers in our group jointly conducted a review and manually reclassified all “signal” and “unknown” components as appropriate. The unique variance from all “noise” components was regressed out, while shared variance was preserved. Following denoising, MSMAll surface registration was performed to enhance anatomical-functional correspondence across participants by integrating multi-modal features, including cortical folding patterns, myelin maps, and resting-state networks ^52^.

### 2.7. RFT Behavioral Analyses

Each participant’s task performance, including accuracy rate and RT for correct trials, was averaged across all RFT runs for each of the 4 cue types (0¢, 10¢, 50¢, and ?). In the full sample, we used ANOVA with *post hoc* pairwise tests to examine the effects of cue type on overall task performance. We then conducted linear regression analyses to assess whether cannabis use and depression were independently associated with cue-related task performance in both the full sample and within the group of adolescents who used cannabis. All analyses were controlled for age and sex.

### 2.8. fMRI Analyses

#### Run-level Analysis

Functional MRI data from the RFT were first spatially smoothed (4 mm FWHM) in CIFTI grayordinate space using Connectome Workbench (Washington University, Missouri, US). The smoothed CIFTI data were then separated into two cortical GIFTI (2D surface) files and one subcortical NIFTI (3D volume) file. Cortical GIFTIs were further split into individual frames (374 frames/run) using the *gifti* and *spm_file* functions of Statistical Parametric Mapping (SPM) v25.01 (Wellcome Trust Centre for Neuroimaging, London, UK) ^53^, based on the 32k midthickness surface meshes for the left and right hemispheres generated during preprocessing. These steps ensured surface-based compatibility with SPM for downstream analyses while preserving signal fidelity in CIFTI space.

#### Participant-level Analysis

Participant-level analyses were conducted in SPM 25.01 using MATLAB R2022a (MathWorks, Massachusetts, US). Consistent with prior work ^27^, the BOLD signal was modeled by convolving the hemodynamic response function with task events. The general linear model (GLM) included 11 regressors: 4 for cue onset, 3 for correct feedback following certain cues, 3 for correct feedback following uncertain cues, and 1 for incorrect (commission error) or too-slow (omission error) feedback. The primary contrasts of interest were: (1) reward expectancy: difference between reward (50¢ + 10¢) cues and non-reward (0¢) cues; and 2) reward attainment: difference between feedback following reward cues and feedback following non-reward cues on correct trials. Neural responses were modeled separately for each participant’s cortical GIFTIs and subcortical NIFTI. The resulting contrast maps were reconstructed back into whole-brain CIFTI format for each participant.

Next, we parcellated participant-level whole brain CIFTI contrast maps using the Cole-Anticevic Brain-wide Network Partition (CAB-NP) ^54^, a high-quality extension of the landmark HCP cortical parcellation ^55^ to segment the entire brain into discrete nodes based on resting-state functional connectivity profiles. As in our prior works ^44,56^, we further subdivided the somatomotor strip based on somatotopic representations released by the HCP to yield a total of 750 whole-brain parcels. This parcellation approach enabled us to balance anatomical interpretability with statistical power. All participant-level parcellated CIFTI contrast maps were concatenated across the full sample for group-level analyses.

#### Group-level Analysis

We conducted group-level analyses using version alpha119 of FSL PALM ^57^, applying non-parametric permutation and sign-flipping procedures to control the family-wise error (FWE) rate. This approach is recommended by neuroimaging statisticians for its strong control of Type I errors and resilience to non-normal or skewed data distributions ^58^. As we aimed to comprehensively assess neural function across the reward system, a network-focused approach was employed. Particularly, reward network masks, comprising the top 10% of nodes activated by reward expectancy and attainment contrasts and corresponding contralateral nodes identified in a prior study ^44^, were applied. The reward expectancy and attainment network masks respectively included 114 and 103 parcels. Parallelized group-level analyses were facilitated using the *PALM-from-Excel* MATLAB script ^59^. Results were considered significant at two-tailed *p_FWE_* < 0.05. Effect sizes of significant results were computed from t-values and degrees of freedom (see **Supplementary Methods**).

In group-level GLMs, the dependent variable was neural response during either reward expectancy or reward attainment. For analyses testing the main effects of cannabis use (first hypothesis) and depression (second hypothesis), the design matrix included cannabis use and depression severity as variables of interest, with age, sex, and head coil type included as covariates of no interest. This approach allowed for assessment of each main effect (cannabis use or depression) independent of the other. If there was a significant main effect, cannabis use × depression interaction effects were subsequently examined with an interaction term (third hypothesis). All regressors were mean-centered, and interaction terms were computed as the products of the mean-centered main variables.

#### Exploratory Analysis

We repeated the parcellated network analyses separately in female-only and male-only subgroups. Within each sex subgroup, models examined neural responses to either reward expectancy or attainment in association with cannabis use and depression, adjusted for age and coil type.

### 2.9. Other Statistics

To statistically describe demographic and clinical characteristics of the study sample, we first assessed the normality and expected frequency for continuous and categorical variables, respectively. Appropriate group comparisons and correlations were subsequently performed. Results were considered significant at *p* < 0.05.

## 3. Results

### 3.1. Demographic and Clinical Characteristics

This study included 117 participants (age: 15.5 ± 2.3 years; 62.4% female). Participants were drawn from a larger sample whose clinical data were previously reported ^30^. Reward flanker task fMRI data from 72 of the participants in the current study were reported in our prior work ^27^. Participants represented a wide range of clinical severity: some met DSM diagnostic criteria for mood, anxiety, or other co-occurring psychiatric disorders; others exhibited subthreshold depressive symptoms; and a portion had neither a past nor current psychiatric diagnosis. Of the 32 participants who used cannabis, 16 reported heavy use (4 met criteria for CUD), 3 reported moderate use, and 13 reported low use. **Table 1** provides details on the demographic and clinical profiles of the sample.

### 3.2. Task Accuracy and Reaction Time

In the full sample, only 4.67 ± 4.83% of the total trials received no response (i.e. omitted trials). The accuracy rate in response to the flanker stimulus across all RFT trials were 86.2 ± 10.0%. There was no significant difference (F_(3,348)_ = 1.52, *p* = 0.209) in accuracy following the four cue types of 0¢ (86.2 ± 13.1%), 10¢ (85.2 ± 11.5%), 50¢ (87.0 ± 11.9%), and(86.3 ± 10.3%). Neither cannabis use nor depression severity showed significant association with cue-specific accuracy.

The overall reaction time (RT) across all trials was 701.7 ± 126.5 ms. A significant difference in RT (F_(3,348)_ = 4.57; Cohen’s *f =* 0.199; *p* = 3.72 × 10^−3^) was observed across cue types: 0¢ (702.1 ± 127.7 ms), 10¢ (695.0 ± 132.5 ms), 50¢ (691.2 ± 130.2 ms), and(706.9 ± 128.6 ms). *Post hoc* pairwise comparisons showed that RT in response to high reward (50¢) trials was significantly faster (Hedges’ *g* = −0.439; *p* = 4.21 × 10^−3^) than in response to uncertain (?) trials. In the full sample, when adjusted for age, sex, and depression severity, adolescents who used cannabis reacted faster than those who did not use cannabis only in high reward trials (β ± SE = −78.3 ± 28.2; *p* = 6.55 × 10^−3^). Within the group of adolescents who used cannabis, cannabis use severity was not significantly associated with RT across cue types. No significant association between depression severity and RT across cue types was detected in either the full sample or the cannabis use group.

### 3.2. Relationships between Cannabis Use and Neural Reward Processing

Across the full sample, there were no significant group differences in neural responses during reward expectancy or attainment between adolescents who used cannabis and those who did not. Among adolescents who used cannabis, heavier cannabis use was associated with greater activity in the mediodorsal and posterior regions of the thalamus during reward expectancy (Figure 2A). Further review using the Morel atlas ^60,61^ specified involvement of the parafascicular thalamic nuclei, nucleus limitans, and habenula.

**Figure 2.**
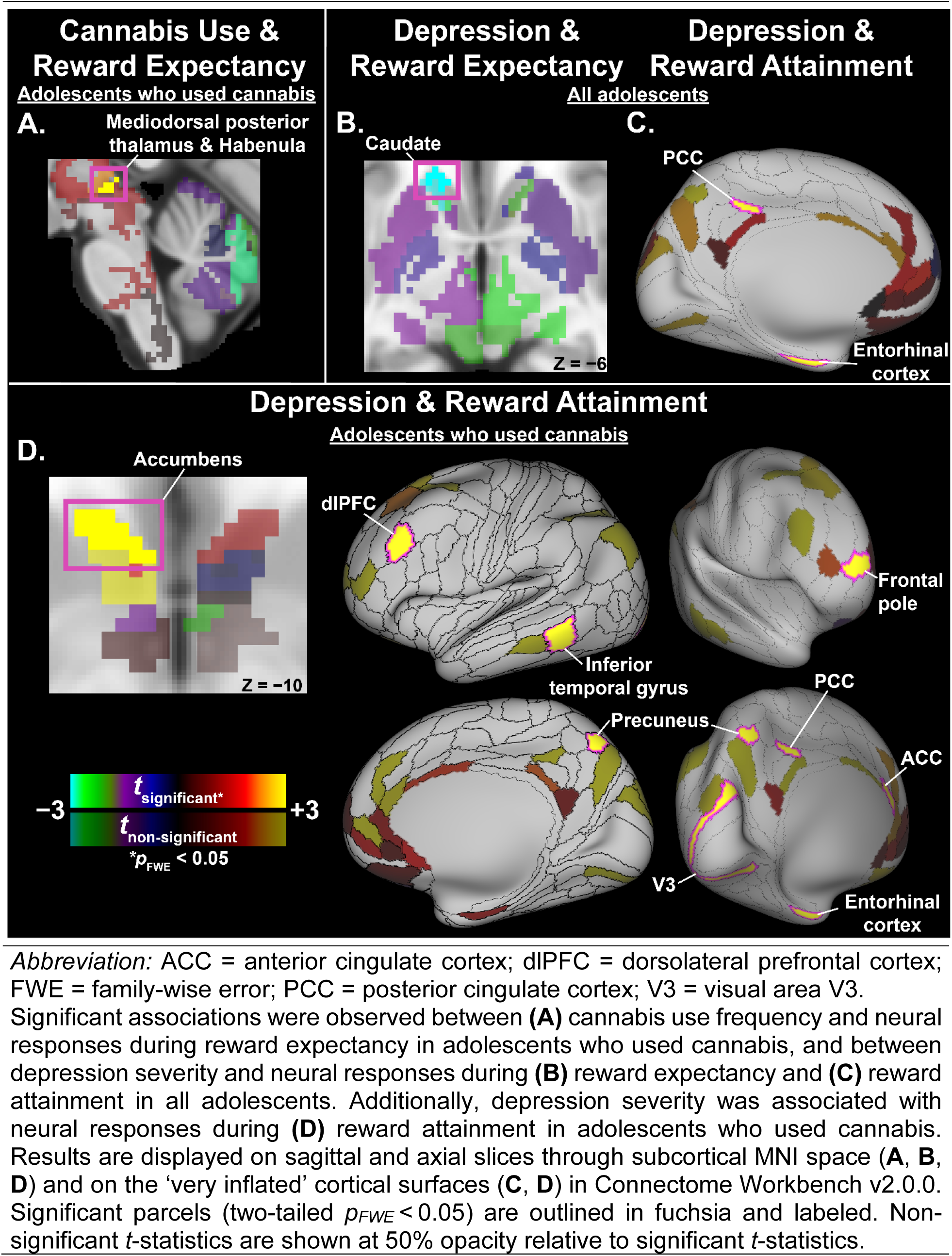
Significant associations between neural responses during reward processes and cannabis use and depression severity.

### 3.3. Relationships between Depression and Neural Reward Processing

Among all participants, more severe depression was linked to blunted activity in the left caudate during reward expectancy (Figure 2B) and elevated activity of the left posterior cingulate cortex (PCC) and entorhinal cortex during reward attainment (Figure 2C). When analyses were restricted to adolescents who used cannabis, more widespread depression-related neural alterations emerged during reward attainment, particularly in reward-related regions overlapping with the default mode, salience, central executive, and visual networks. Specifically, greater depression severity was associated with hyperactivation in the left PCC, entorhinal cortex, dorsolateral prefrontal cortex (dlPFC), inferior temporal gyrus, visual cortex area V3, anterior cingulate cortex (ACC), nucleus accumbens, right frontal pole, and bilateral precuneus (Figure 2D).

### 3.4. Interaction of Cannabis Use and Depression in Reward Processing

There was a significant cannabis use × depression interaction effect on task-evoked activation during reward attainment. In the full sample, the interaction between cannabis use and depression severity was significantly associated with hyperactivation in the left inferior temporal gyrus and frontal pole (Figure 3A). Among adolescents who used cannabis, a positive interaction between cannabis use frequency and depression severity emerged in the left entorhinal cortex (Figure 3B). In contrast, no interaction effect was detected during reward expectancy. **Table 2** details all significant findings described in sections 3.3 – 3.5.

**Figure 3.**
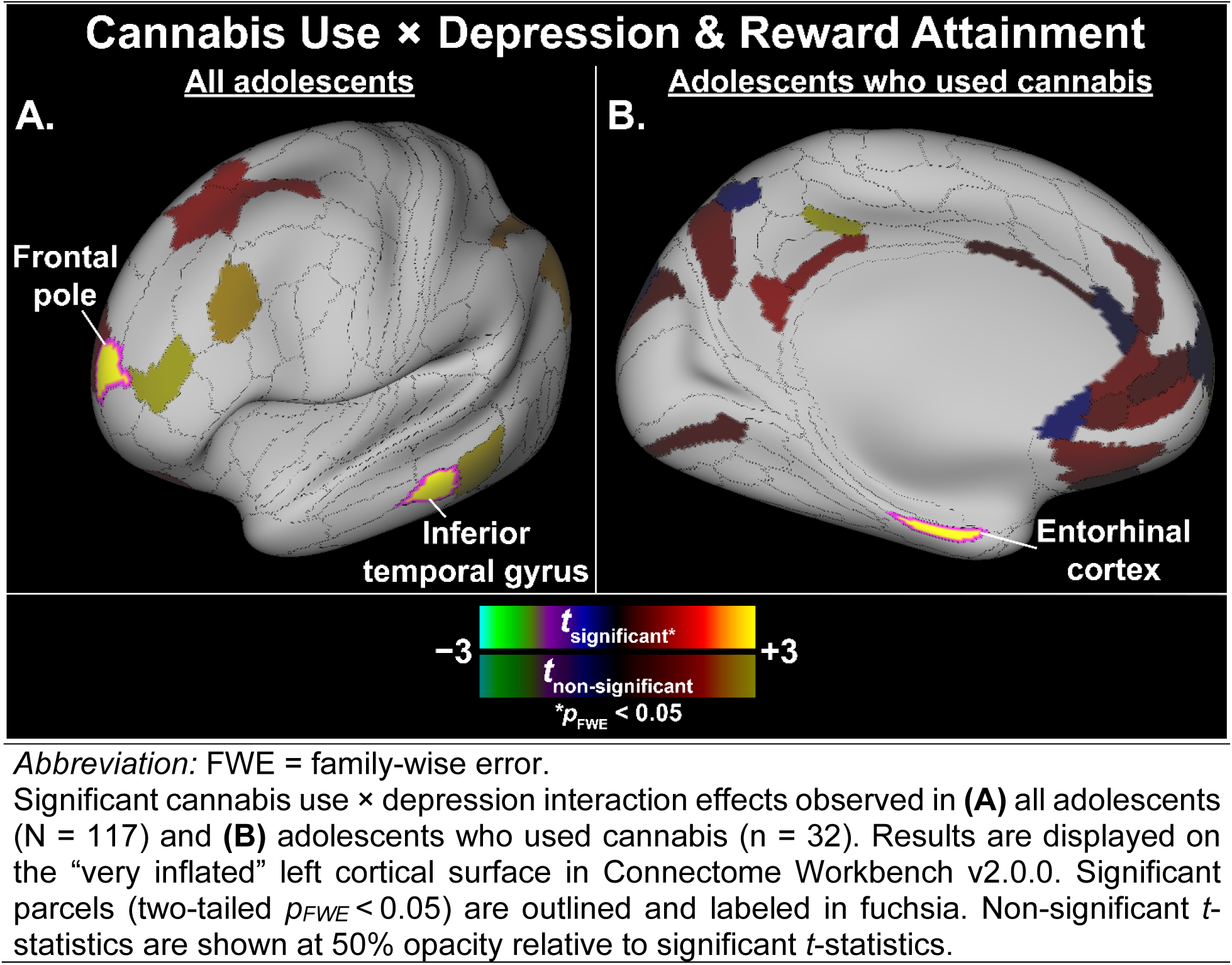
Significant cannabis use × depression interaction effect on neural responses during reward attainment.

**Table 2.** Significant associations between cannabis use, depression, and neural responses during reward expectancy and attainment.

| RFT Contrast | Region (Lateralization) | HCP Cortical Label<sup>a</sup> | CAB-NP Label<sup>b</sup> | t-statistic<sup>c</sup> | Pearson's r<sup>d</sup> | p<sub>FWE</sub> |
| --- | --- | --- | --- | --- | --- | --- |
| **All Adolescents (N = 117) – Depression Severity<sup>e</sup>** |  |  |  |  |  |  |
| Reward Expectancy | Caudate (L) | — | Visual-13 | -3.79 | -0.340 | 0.0209 |
| Reward Attainment | Dorsal posterior cingulate cortex (L) | 31a | Default_72 | 4.09 | 0.363 | 7.60 × 10<sup>-3</sup> |
|  | Entorhinal cortex (L) | EC | Default_60 | 3.58 | 0.323 | 0.0411 |
| **Adolescents with Cannabis Use (n = 32) – Cannabis Use Frequency<sup>f</sup>** |  |  |  |  |  |  |
| Reward Expectancy | Mediodorsal posterior thalamus/habenula (L) | — | Visual-58 | 4.48 | 0.660 | 0.0113 |
| **Adolescents with Cannabis Use (n = 32) – Depression Severity<sup>e</sup>** |  |  |  |  |  |  |
| Reward Attainment | Dorsolateral prefrontal cortex (L) | p9-46v | Frontoparietal_36 | 5.70 | 0.746 | 1.00 × 10<sup>-3</sup> |
|  | Inferior temporal gyrus (L) | TE1p | Frontoparietal_44 | 4.67 | 0.675 | 8.50 × 10<sup>-3</sup> |
|  | Visual cortex (L) | V3 | Visual2_30 | 3.95 | 0.612 | 0.0448 |
|  | Frontal pole (R) | p10p | Frontoparietal_25 | 4.11 | 0.628 | 0.0326 |
|  | Anterior cingulate cortex (L) | p24 | Cingulo-Opercular_55 | 3.97 | 0.614 | 0.0433 |
|  | Entorhinal cortex (L) | EC | Default_60 | 5.01 | 0.701 | 3.50 × 10<sup>-3</sup> |
|  | Dorsal posterior cingulate (L) | 31a | Default_72 | 4.09 | 0.625 | 0.0144 |
|  | Precuneus (L) | 7Pm | Frontoparietal_30 | 4.54 | 0.665 | 0.0112 |
|  | Precuneus (R) | 7Pm | Frontoparietal_2 | 4.54 | 0.665 | 0.0113 |
|  | Accumbens (L) | — | Visual-2 | 4.29 | 0.644 | 0.0212 |
| **All Adolescents (N = 117) – Cannabis Use × Depression Interaction** |  |  |  |  |  |  |
| Reward Attainment | Inferior temporal gyrus (L) | TE1m | Default_76 | 3.72 | 0.335 | 0.0313 |
|  | Frontal pole (L) | p10p | Frontoparietal_48 | 3.94 | 0.353 | 0.0141 |
| **Adolescents with Cannabis Use (n = 32) – Cannabis Use × Depression Interaction** |  |  |  |  |  |  |
| Reward Attainment | Entorhinal cortex (L) | EC | Default_60 | 3.96 | 0.621 | 0.0482 |

**Abbreviations:** CAB-NP = Cole-Anticevic Brain-wide Network Partition; FWE = Family-wise error; HCP = Human Connectome Project; L = Left; R = Right; RFT = Reward Flanker Task.
<sup>a</sup> Labels per the HCP Cortical Atlas, as detailed in Glasser et al. (2016).
<sup>b</sup> Labels per Cole-Anticevic Brain-wide Network Partition v1.0.5 (equivalent labels per HCP S1200 Release cortical parcellation), which comprised 392 cortical nodes derived from HCP parcellation and 358 subcortical nodes from Cole-Anticevic atlas.
<sup>c</sup> Adjusted for age, sex, and head coil type.
<sup>d</sup> Pearson's *r* represents effect size and was computed from *t*-statistic and its corresponding degree of freedom output from FSL PALM.
<sup>e</sup> Depression effect was adjusted for cannabis use effect.
<sup>f</sup> Cannabis use effect was adjusted for depression effect.

### 3.5. Exploratory Results on Sex Differences

Among female adolescents (n = 73), 31.5% reported cannabis use (n = 23; 11 with heavy use, 3 with moderate use, and 9 low use). Among male adolescents (n = 44), 20.5% reported cannabis use (n = 9; 5 with heavy use and 4 with low use). There was no statistical difference (p = 0.194) in the proportion of those with cannabis use between female and male groups. Depression severity also did not differ significantly (p = 0.445) between female (CDSR-R: 34.9 ± 14.7) and male (CDSR-R: 33.8 ± 16.7) participants.

#### Female-only results

Activation of the left mediodorsal thalamus and habenula during reward expectancy was associated with more frequent cannabis use among female adolescents who used cannabis, consistent with the primary finding. Only activation in the left inferior temporal gyrus during reward attainment was significantly associated with depression severity in female adolescents who used cannabis.

#### Male-only results

Across all male participants, depression severity was negatively correlated with activation in the left caudate during reward expectancy.

**Supplementary Figure S1** and **Table S5** detail sex-specific results.

## 4. Discussion

In the present study, we examined neural function underlying reward expectancy and attainment in relation to comorbid adolescent cannabis use and depression. Using a parcellated, network-focused approach, we assessed cortical and subcortical regions across the reward circuitry, thereby complementing and extending prior adolescent fMRI studies focusing on regions of interest ^17,62^. Supporting our hypotheses, cannabis use was associated with altered neural responses during reward expectancy, but not attainment. Depression severity was linked to disruptions across both reward phases, with more widespread alterations during reward attainment among adolescents who used cannabis. There was a significant interaction effect between cannabis use and depression on neural activity during reward attainment. Contrary to our hypothesis, no interaction effect was observed during reward expectancy. Exploratory analyses further revealed sex-specific findings on reward dysfunction in this comorbidity. We discuss our findings in detail below.

### 4.1. Reward Dysfunction in Adolescent Cannabis Use and Depression

Behaviorally, adolescents who used cannabis responded faster to flanker stimuli following high reward cues than those who did not. This is in contrast with the commonly observed slower processing in adolescent cannabis use across various cognitive domains ^63^, likely explained by our reward-focused task design. Indeed, faster responses without compromising accuracy may reflect heightened sensitivity to high reward cues in cannabis-using youth. Our results also differ from prior fMRI studies using the Monetary Incentive Delay (MID) reward task ^24^, which have reported minimal associations between adolescent cannabis use and behavioral performance ^17^. This discrepancy underscores the importance of separating cue valence while engaging cognitive control when assessing reward function in adolescence, a period marked by earlier maturation of motivational systems relative to top-down regulatory mechanisms ^23^.

During reward expectancy, although neural responses did not differ between those with and without cannabis use, heavier use within the cannabis use group was associated with hyperactivation in a parcel overlapping with the mediodorsal thalamus, including the parafascicular nucleus and the habenula. As CAB-NP parcels are defined by functional connectivity ^54^, these anatomical references indicate overlap rather than precise localization. Nonetheless, these regions have not been previously implicated in cannabis-related reward expectancy in fMRI studies ^17^. Preclinical work suggests that the mediodorsal thalamus coordinates signaling with cortical areas for reward valuation and action preparation ^64,65^, while the parafascicular nucleus, projecting to the nucleus accumbens ^66^, supports action initiation ^67^. The habenula is a core node in the anti-reward system, modulating dopaminergic tone and encoding aversive predictions ^68^. Thus, adolescents with heavier cannabis use may exhibit altered valuation, undervaluing rewards even while preparing to exert effort to obtain them. These seemingly paradoxical processes are possibly driven by cannabinoid-induced downregulation of CB_1_ receptors densely expressed in the parafascicular thalamus and habenula ^69,70^, leading to reduced presynaptic inhibition and heightened activity. Further, these neural impairments appeared more pronounced in female adolescents, extending prior evidence of greater female susceptibility to cannabis-related neurodevelopmental changes ^5,18^ by specifying vulnerability in reward expectancy. More broadly, our findings offer further insight into the neural basis of motivational deficits in frequent cannabis use ^71^.

Independent of cannabis use, greater depression severity was associated with altered neural responses during both reward expectancy and attainment, consistent with prior findings ^19–21^. During expectancy, depression-linked blunted caudate activity was observed across the full sample but was non-significant in cannabis-using adolescents, suggesting that cannabinoid effects may dominate co-occurring depression effects on expectancy. Moreover, sex-stratified analyses showed that this depression-related caudate attenuation during expectancy was significant in males but not females. Though sex-stratified analyses were exploratory, this is surprising given the female propensity for depression, but could be explained by a lag in striatal development in boys vs. girls ^72^.

During reward attainment, greater depression severity was associated with increased activation in the PCC and entorhinal cortex across the full sample. Hyperactivity of the PCC, and the default network more broadly, reflects heightened self-referential processing and has been linked to rumination in depression ^73^. The entorhinal cortex is a key navigation center involved in memory encoding and can aid reward learning based on emerging preclinical evidence ^74^, yet it has rarely been implicated in depression. Among adolescents with cannabis use, greater depression severity further involved the nucleus accumbens, ACC, dlPFC, precuneus, and visual cortex regions. One integrated interpretation is that, when receiving rewards after expending cognitive effort (as in our task), adolescents with cannabis use and more severe depression may experience reward outcomes as more salient, prompting excessive self-reflection and memory processing. This may imply altered reward learning. In addition, we observed an interaction effect during reward attainment in which cannabis use intensified the relationship between depression severity and activation of the frontal pole, inferior temporal gyrus, and entorhinal cortex. This supports an additive effect of cannabis use and depression on reward function, particularly in regions that are part of executive control and memory systems. Taken together, our results demonstrate that neural reward dysfunction in concurrent cannabis use and depression differs from that in either condition alone, emphasizing the need to investigate shared and differential neural mechanisms underlying co-occurring psychopathology.

### 4.2. Clinical Implications

Our results provide translational implications for clinical management of adolescents with co-occurring cannabis use and depression. In line with our fMRI results, interventions that target reward, self-reflection, and executive control processes may be particularly well-suited for this comorbidity. Embedding neurobiologically informed education in school and family settings can further improve treatment adherence and cessation by highlighting the impact of cannabis on brain development and health outcomes ^75^. As optimizing depression treatment can reduce cannabis use ^76^, and cannabis use reduction may improve depression trajectory ^77^, early detection and integrated management of both conditions is essential.

### 4.3. Study Limitations and Future Directions

Our study has several limitations. First, its cross-sectional design precludes causal interpretations. Follow-up longitudinal research is needed to delineate the consequences of cannabis use and depression on reward system development and clinical trajectories. Second, cannabis use patterns were primarily characterized through retrospective self-reports, which are susceptible to recall and social desirability biases. Biological measures of cannabinoids should be incorporated in subsequent studies to precisely capture dose-dependent associations with reward dysfunction. Future studies should also explore the role of social rewards, given their relevance to both conditions in adolescence ^78,79^. Third, although the group size of adolescents with cannabis use was modest, it was similar to other reward task fMRI studies on cannabis use in youth ^17^. *Post hoc* sensitivity analyses further indicated that our study achieved 80% power to detect effects as small as |*r*| = 0.25 in the full sample, |*r*| = 0.46 in the group of adolescents who used cannabis, and *d* = 0.59 for group comparisons between adolescents with vs. without cannabis use. The proportion of adolescents endorsing cannabis use in our sample (∼27%) also resembles general population estimates ^11,80^, bolstering the generalizability of our findings. Additionally, our ability to detect neurobiological correlates of adolescent cannabis use and depression was enhanced by our methodological approach, including high-quality MRI acquisitions and sophisticated surface-based (CIFTI) analyses for enhanced inter-subject alignment and signal localization ^81^; these advanced techniques remain underutilized in pediatric neuroimaging.

### 4.4. Conclusions

Our investigation of adolescent depression and cannabis use comorbidity integrated a reward fMRI task with rigorous acquisition and analytic methods to study this high-risk population. We identified distinct neural profiles of reward processing alterations related to cannabis use, depression, and their interaction. As suggested by these findings, interventions targeting both anticipatory and consummatory aspects of reward function should be implemented for concurrent cannabis use and depression in youth. Such efforts are critical given the escalating potency and accessibility of cannabinoid products, which pose substantial risks to reward systems during a vulnerable developmental period.

## Data Availability

All data produced in the present study are available upon reasonable request to the corresponding author.

## References

1. Centers for Disease, C. & Prevention. (2021).

2 Hughes, A., Lipari, R. N. & Williams, M. The CBHSQ Report: State Estimates of Adolescent Marijuana Use and Perceptions of Risk of Harm from Marijuana Use: 2013 and 2014. Substance Abuse and Mental Health Services Administration, Center for Behavioral Health Statistics and Quality (2015).

3 Mital, S. & Nguyen, H. V. Legalizing Youth-Friendly Cannabis Edibles and Extracts and Adolescent Cannabis Use. JAMA Netw Open 8, e255819 (2025). 10.1001/jamanetworkopen.2025.5819

4 Galván, A. The Teenage Brain. Current Directions in Psychological Science 22, 88–93 (2013). 10.1177/0963721413480859

5 Meyer, H. C., Lee, F. S. & Gee, D. G. The Role of the Endocannabinoid System and Genetic Variation in Adolescent Brain Development. Neuropsychopharmacology 43, 21–33 (2018). 10.1038/npp.2017.143

6 Tseng, K. Y. & Molla, H. M. Cannabinoid CB1 receptor-sensitive neurodevelopmental processes and trajectories. Mol Psychiatry 30, 3792–3803 (2025). 10.1038/s41380-025-03057-2

7 Parsons, L. H. & Hurd, Y. L. Endocannabinoid signalling in reward and addiction. Nat Rev Neurosci 16, 579–594 (2015). 10.1038/nrn4004

8 Miller, M. L. et al. Adolescent exposure to Delta(9)-tetrahydrocannabinol alters the transcriptional trajectory and dendritic architecture of prefrontal pyramidal neurons. Mol Psychiatry 24, 588–600 (2019). 10.1038/s41380-018-0243-x

9 Wenzel, J. M. & Cheer, J. F. Endocannabinoid Regulation of Reward and Reinforcement through Interaction with Dopamine and Endogenous Opioid Signaling. Neuropsychopharmacology 43, 103–115 (2018). 10.1038/npp.2017.126

10 Gobbi, G. et al. Association of Cannabis Use in Adolescence and Risk of Depression, Anxiety, and Suicidality in Young Adulthood: A Systematic Review and Meta-analysis. JAMA Psychiatry 76, 426–434 (2019). 10.1001/jamapsychiatry.2018.4500

11 Sultan, R. S., Zhang, A. W., Olfson, M., Kwizera, M. H. & Levin, F. R. Nondisordered Cannabis Use Among US Adolescents. JAMA Netw Open 6, e2311294 (2023). 10.1001/jamanetworkopen.2023.11294

12 Wilkinson, A. L. et al. Testing longitudinal relationships between binge drinking, marijuana use, and depressive symptoms and moderation by sex. Journal of Adolescent Health 59, 681–687 (2016). 10.1016/j.jadohealth.2016.07.010

13 Bataineh, B. S. et al. The Association Between Tobacco and Cannabis Use and the Age of Onset of Depression and Anxiety Symptoms: Among Adolescents and Young Adults. Nicotine & tobacco research : official journal of the Society for Research on Nicotine and Tobacco 25, 1455–1464 (2023). 10.1093/ntr/ntad058

14 Weinberger, A. H. et al. Cannabis use among youth in the United States, 2004-2016: Faster rate of increase among youth with depression. Drug Alcohol Depend 209, 107894 (2020). 10.1016/j.drugalcdep.2020.107894

15 Marel, C. et al. Conditional probabilities of substance use disorders and associated risk factors: Progression from first use to use disorder on alcohol, cannabis, stimulants, sedatives and opioids. Drug Alcohol Depend 194, 136–142 (2019). 10.1016/j.drugalcdep.2018.10.010

16 Groenman, A. P., Janssen, T. W. P. & Oosterlaan, J. Childhood Psychiatric Disorders as Risk Factor for Subsequent Substance Abuse: A Meta-Analysis. J Am Acad Child Adolesc Psychiatry 56, 556–569 (2017). 10.1016/j.jaac.2017.05.004

17 Beyer, E., Poudel, G., Antonopoulos, S., Thomson, H. & Lorenzetti, V. Brain reward function in people who use cannabis: a systematic review. Front Behav Neurosci 17, 1323609 (2023). 10.3389/fnbeh.2023.1323609

18 Hammond, C. J. et al. A Meta-Analysis of fMRI Studies of Youth Cannabis Use: Alterations in Executive Control, Social Cognition/Emotion Processing, and Reward Processing in Cannabis Using Youth. Brain Sci 12 (2022). 10.3390/brainsci12101281

19 Forbes, E. E. et al. Altered striatal activation predicting real-world positive affect in adolescent major depressive disorder. Am J Psychiatry 166, 64–73 (2009). 10.1176/appi.ajp.2008.07081336

20 Keren, H. et al. Reward Processing in Depression: A Conceptual and Meta-Analytic Review Across fMRI and EEG Studies. Am J Psychiatry 175, 1111–1120 (2018). 10.1176/appi.ajp.2018.17101124

21 Rzepa, E., Fisk, J. & McCabe, C. Blunted neural response to anticipation, effort and consummation of reward and aversion in adolescents with depression symptomatology. J Psychopharmacol 31, 303–311 (2017). 10.1177/0269881116681416

22 Richards, J. M., Plate, R. C. & Ernst, M. A systematic review of fMRI reward paradigms used in studies of adolescents vs. adults: the impact of task design and implications for understanding neurodevelopment. Neurosci Biobehav Rev 37, 976–991 (2013). 10.1016/j.neubiorev.2013.03.004

23 Shulman, E. P. et al. The dual systems model: Review, reappraisal, and reaffirmation. Dev Cogn Neurosci 17, 103–117 (2016). 10.1016/j.dcn.2015.12.010

24 Knutson, B., Adams, C. M., Fong, G. W. & Hommer, D. Anticipation of increasing monetary reward selectively recruits nucleus accumbens. J Neurosci 21, RC159 (2001). 10.1523/JNEUROSCI.21-16-j0002.2001

25 Eriksen, B. A. & Eriksen, C. W. Effects of noise letters upon the identification of a target letter in a nonsearch task. Perception & Psychophysics 16, 143–149 (1974). 10.3758/bf03203267

26 Bradley, K. A. L., Case, J. A. C., Freed, R. D., Stern, E. R. & Gabbay, V. Neural correlates of RDoC reward constructs in adolescents with diverse psychiatric symptoms: A Reward Flanker Task pilot study. J Affect Disord 216, 36–45 (2017). 10.1016/j.jad.2016.11.042

27 Liu, Q. et al. Neural function underlying reward expectancy and attainment in adolescents with diverse psychiatric symptoms. Neuroimage Clin 36, 103258 (2022). 10.1016/j.nicl.2022.103258

28 Ely, B. A., Nguyen, T. N. B., Tobe, R. H., Walker, A. M. & Gabbay, V. Multimodal Investigations of Reward Circuitry and Anhedonia in Adolescent Depression. Front Psychiatry 12, 678709 (2021). 10.3389/fpsyt.2021.678709

29 Treadway, M. T. & Zald, D. H. Reconsidering anhedonia in depression: lessons from translational neuroscience. Neurosci Biobehav Rev 35, 537–555 (2011). 10.1016/j.neubiorev.2010.06.006

30 Nguyen, T. N. B. et al. Cannabis Use is Related to Anhedonia in Adolescents With Diverse Mood and Anxiety Symptoms. JAACAP Open (2025). 10.1016/j.jaacop.2025.02.003

31 Mohammadi, S. et al. Brain-based Sex Differences in Depression: A Systematic Review of Neuroimaging Studies. Brain Imaging Behav 17, 541–569 (2023). 10.1007/s11682-023-00772-8

32 Kaufman, J. et al. Schedule for Affective Disorders and Schizophrenia for School-Age Children-Present and Lifetime Version (K-SADS-PL): initial reliability and validity data. J Am Acad Child Adolesc Psychiatry 36, 980–988 (1997). 10.1097/00004583-199707000-00021

33 Sheehan, D. V. et al. The Mini-International Neuropsychiatric Interview (M.I.N.I.): the development and validation of a structured diagnostic psychiatric interview for DSM-IV and ICD-10. J Clin Psychiatry 59 Suppl 20, 22–33;quiz 34-57 (1998).

34 Smith, D. K. Kaufman Brief Intelligence Test (K-BIT). Diagnostique 24, 125–134 (1999). 10.1177/153450849902401-412

35 Hudziak, J. J., Achenbach, T. M., Althoff, R. R. & Pine, D. S. A dimensional approach to developmental psychopathology. Int J Methods Psychiatr Res 16 Suppl 1, S16–23 (2007). 10.1002/mpr.217

36 Poznanski, E. O. & Mokros, H. B. Children’s Depression Rating Scale–Revised (CDRS-R). (WPS, 1996).

37 Mayes, T. L., Bernstein, I. H., Haley, C. L., Kennard, B. D. & Emslie, G. J. Psychometric properties of the Children’s Depression Rating Scale-Revised in adolescents. J Child Adolesc Psychopharmacol 20, 513–516 (2010). 10.1089/cap.2010.0063

38 March, J. S., Parker, J. D., Sullivan, K., Stallings, P. & Conners, C. K. The Multidimensional Anxiety Scale for Children (MASC): factor structure, reliability, and validity. J Am Acad Child Adolesc Psychiatry 36, 554–565 (1997). 10.1097/00004583-199704000-00019

39 Beck, A. T., Kovacs, M. & Weissman, A. Assessment of suicidal intention: the Scale for Suicide Ideation. J Consult Clin Psychol 47, 343–352 (1979). 10.1037//0022-006x.47.2.343

40 Gard, D. E., Gard, M. G., Kring, A. M. & John, O. P. Anticipatory and consummatory components of the experience of pleasure: a scale development study. Journal of research in personality 40, 1086–1102 (2006).

41 Harms, M. P. et al. Extending the Human Connectome Project across ages: Imaging protocols for the Lifespan Development and Aging projects. Neuroimage 183, 972–984 (2018). 10.1016/j.neuroimage.2018.09.060

42 Liu, Q. et al. Correlates of C-reactive protein with neural reward circuitry in adolescents with psychiatric symptoms. Brain Behav Immun Health 9 (2020). 10.1016/j.bbih.2020.100153

43 Liu, Q. et al. Reward function as an outcome predictor in youth with mood and anxiety symptoms. J Affect Disord 278, 433–442 (2021). 10.1016/j.jad.2020.09.074

44 Ely, B. A. et al. Data-driven parcellation and graph theory analyses to study adolescent mood and anxiety symptoms. Transl Psychiatry 11, 266 (2021). 10.1038/s41398-021-01321-x

45 Morris, L. S. et al. Ketamine normalizes subgenual cingulate cortex hyper-activity in depression. Neuropsychopharmacology 45, 975–981 (2020). 10.1038/s41386-019-0591-5

46 Costi, S. et al. Impact of the KCNQ2/3 Channel Opener Ezogabine on Reward Circuit Activity and Clinical Symptoms in Depression: Results From a Randomized Controlled Trial. Am J Psychiatry 178, 437–446 (2021). 10.1176/appi.ajp.2020.20050653

47 de Bie, H. M. et al. Preparing children with a mock scanner training protocol results in high quality structural and functional MRI scans. Eur J Pediatr 169, 1079–1085 (2010). 10.1007/s00431-010-1181-z

48 Glasser, M. F. et al. The minimal preprocessing pipelines for the Human Connectome Project. Neuroimage 80, 105–124 (2013). 10.1016/j.neuroimage.2013.04.127

49 Grabner, G. et al. Symmetric atlasing and model based segmentation: an application to the hippocampus in older adults. Med Image Comput Comput Assist Interv 9, 58–66 (2006). 10.1007/11866763_8

50 Griffanti, L. et al. ICA-based artefact removal and accelerated fMRI acquisition for improved resting state network imaging. Neuroimage 95, 232–247 (2014). 10.1016/j.neuroimage.2014.03.034

51 Glasser, M. F. et al. Using temporal ICA to selectively remove global noise while preserving global signal in functional MRI data. Neuroimage 181, 692–717 (2018). 10.1016/j.neuroimage.2018.04.076

52 Robinson, E. C. et al. MSM: a new flexible framework for Multimodal Surface Matching. Neuroimage 100, 414–426 (2014). 10.1016/j.neuroimage.2014.05.069

53 Tierney, T. M. et al. SPM 25: open source neuroimaging analysis software. Journal of Open Source Software 10 (2025). 10.21105/joss.08103

54 Ji, J. L. et al. Mapping the human brain’s cortical-subcortical functional network organization. Neuroimage 185, 35–57 (2019). 10.1016/j.neuroimage.2018.10.006

55 Glasser, M. F. et al. A multi-modal parcellation of human cerebral cortex. Nature 536, 171–178 (2016). 10.1038/nature18933

56 Ely, B. A., Stern, E. R., Kim, J. W., Gabbay, V. & Xu, J. Detailed mapping of human habenula resting-state functional connectivity. Neuroimage 200, 621–634 (2019). 10.1016/j.neuroimage.2019.06.015

57 Winkler, A. M., Ridgway, G. R., Webster, M. A., Smith, S. M. & Nichols, T. E. Permutation inference for the general linear model. Neuroimage 92, 381–397 (2014). 10.1016/j.neuroimage.2014.01.060

58 Winkler, A. M. et al. Non-parametric combination and related permutation tests for neuroimaging. Hum Brain Mapp 37, 1486–1511 (2016). 10.1002/hbm.23115

59 Palm-From-Excel (MathWorks File Exchange, 2024).

60 Krauth, A. et al. A mean three-dimensional atlas of the human thalamus: generation from multiple histological data. Neuroimage 49, 2053–2062 (2010). 10.1016/j.neuroimage.2009.10.042

61 Jakab, A., Blanc, R., Berenyi, E. L. & Szekely, G. Generation of individualized thalamus target maps by using statistical shape models and thalamocortical tractography. AJNR Am J Neuroradiol 33, 2110–2116 (2012). 10.3174/ajnr.A3140

62 O’Callaghan, G. & Stringaris, A. Reward Processing in Adolescent Depression Across Neuroimaging Modalities. Z Kinder Jugendpsychiatr Psychother 47, 535–541 (2019). 10.1024/1422-4917/a000663

63 Scott, J. C. et al. Association of Cannabis With Cognitive Functioning in Adolescents and Young Adults: A Systematic Review and Meta-analysis. JAMA Psychiatry 75, 585–595 (2018). 10.1001/jamapsychiatry.2018.0335

64 Wicker, E., Turchi, J., Malkova, L. & Forcelli, P. A. Mediodorsal thalamus is required for discrete phases of goal-directed behavior in macaques. Elife 7 (2018). 10.7554/eLife.37325

65 Pergola, G. et al. The Regulatory Role of the Human Mediodorsal Thalamus. Trends Cogn Sci 22, 1011–1025 (2018). 10.1016/j.tics.2018.08.006

66 Mandelbaum, G. et al. Distinct Cortical-Thalamic-Striatal Circuits through the Parafascicular Nucleus. Neuron 102, 636–652 e637 (2019). 10.1016/j.neuron.2019.02.035

67 Fallon, I. P. et al. The role of the parafascicular thalamic nucleus in action initiation and steering. Curr Biol 33, 2941–2951 e2944 (2023). 10.1016/j.cub.2023.06.025

68 Hikosaka, O. The habenula: from stress evasion to value-based decision-making. Nat Rev Neurosci 11, 503–513 (2010). 10.1038/nrn2866

69 Tsou, K., Brown, S., Sanudo-Pena, M. C., Mackie, K. & Walker, J. M. Immunohistochemical distribution of cannabinoid CB1 receptors in the rat central nervous system. Neuroscience 83, 393–411 (1998). 10.1016/s0306-4522(97)00436-3

70 Berger, A. L. et al. The Lateral Habenula Directs Coping Styles Under Conditions of Stress via Recruitment of the Endocannabinoid System. Biol Psychiatry 84, 611–623 (2018). 10.1016/j.biopsych.2018.04.018

71 Lac, A. & Luk, J. W. Testing the Amotivational Syndrome: Marijuana Use Longitudinally Predicts Lower Self-Efficacy Even After Controlling for Demographics, Personality, and Alcohol and Cigarette Use. Prev Sci 19, 117–126 (2018). 10.1007/s11121-017-0811-3

72 Lenroot, R. K. et al. Sexual dimorphism of brain developmental trajectories during childhood and adolescence. Neuroimage 36, 1065–1073 (2007). 10.1016/j.neuroimage.2007.03.053

73 Zhou, H. X. et al. Rumination and the default mode network: Meta-analysis of brain imaging studies and implications for depression. Neuroimage 206, 116287 (2020). 10.1016/j.neuroimage.2019.116287

74 Sosa, M. & Giocomo, L. M. Navigating for reward. Nat Rev Neurosci 22, 472–487 (2021). 10.1038/s41583-021-00479-z

75 Steele, D. W. et al. Brief Behavioral Interventions for Substance Use in Adolescents: A Meta-analysis. Pediatrics 146 (2020). 10.1542/peds.2020-0351

76 Curry, J. F. et al. Adaptive Treatment for Youth With Substance Use and Depression: Early Depression Response and Short-term Outcomes. J Am Acad Child Adolesc Psychiatry 61, 508–519 (2022). 10.1016/j.jaac.2021.07.807

77 Hser, Y. I. et al. Reductions in cannabis use are associated with improvements in anxiety, depression, and sleep quality, but not quality of life. J Subst Abuse Treat 81, 53–58 (2017). 10.1016/j.jsat.2017.07.012

78 Forbes, E. E. & Dahl, R. E. Research Review: altered reward function in adolescent depression: what, when and how? J Child Psychol Psychiatry 53, 3–15 (2012). 10.1111/j.1469-7610.2011.02477.x

79 Caouette, J. D. & Feldstein Ewing, S. W. Four Mechanistic Models of Peer Influence on Adolescent Cannabis Use. Curr Addict Rep 4, 90–99 (2017). 10.1007/s40429-017-0144-0

80 Miech, R. A., Johnston, L. D., Patrick, M. E. & O’Malley, P. M. Monitoring the Future national survey results on drug use, 1975–2024: Overview and detailed results for secondary school students. (Institute for Social Research, University of Michigan, 2025).

81 Coalson, T. S., Van Essen, D. C. & Glasser, M. F. The impact of traditional neuroimaging methods on the spatial localization of cortical areas. Proc Natl Acad Sci U S A 115, E6356–E6365 (2018). 10.1073/pnas.1801582115

