## Supplementary Materials for "Neural Correlates of Reward Dysfunction in Adolescent Cannabis Use and Depression"

### Abbreviations

ACC: anterior cingulate cortex

AC-PC: anterior commissure–posterior commissure

ADHD: attention-deficit/hyperactivity disorder

ANOVA: analysis of variance

BDI: Beck Depression Inventory

BOLD: blood oxygen–level dependent

BSSI: Beck Scale for Suicide Ideation

CAB-NP: Cole–Anticevic Brain-wide Network Partition

CDRS-R: Children's Depression Rating Scale–Revised

CIFTI: Connectivity Informatics Technology Initiative

CUD: cannabis use disorder

dlPFC: dorsolateral prefrontal cortex

DSM: Diagnostic and Statistical Manual of Mental Disorders

DSM-5: Diagnostic and Statistical Manual of Mental Disorders, Fifth Edition

EPI: echo-planar imaging

FOV: field of view

FSL: FMRIB Software Library

fMRI: functional magnetic resonance imaging

FWE: family-wise error

GIFTI: Geometry format of the Neuroimaging Informatics Technology Initiative

GLM: generalized linear model

HCP: Human Connectome Project

ICA: independent components analysis

ICA-FIX: ICA-based X-noiseifier

IRB: Institutional Review Boards

IQ: intelligence quotient

K-BIT: Kaufman Brief Intelligence Test

K-SADS-PL: Kiddie Schedule for Affective Disorders and Schizophrenia – Present and Lifetime Version for Children

LR: left-to-right (phase-encoding direction)

MASC: Multidimensional Anxiety Scale for Children

MDMA: 3,4-methylenedioxymethamphetamine

MID: Monetary Incentive Delay

MNI: Montreal Neurological Institute

MPRAGE: magnetization-prepared rapid gradient-echo

MRI: magnetic resonance imaging

MSMAll: multimodal surface matching (“All” variant)

NIFTI: Neuroimaging Informatics Technology Initiative (file format)

PALM: Permutation Analysis of Linear Models

PCC: posterior cingulate cortex

PTSD: post-traumatic stress disorder

RFT: Reward Flanker Task

RL: right-to-left (phase-encoding direction)

ROI: region of interest

RT: reaction time

SD: standard deviation

SPACE: Sampling Perfection with Application optimized Contrast using different flip angle Evolution

SPM: Statistical Parametric Mapping

TE: echo time

TEPS: Temporal Experience of Pleasure Scale

TEPS-A: Temporal Experience of Pleasure Scale – Anticipatory subscale

TEPS-C: Temporal Experience of Pleasure Scale – Consummatory subscale

THC: $\Delta^9$-tetrahydrocannabinol

TR: repetition time

V3: visual area V3

### Supplementary Methods

*SM1. Cannabis Use Quantification*

As described in our prior publication[1] and summarized in **Supplementary Table S1**, participants who did not use cannabis all had negative urine toxicology tests and reported either never using cannabis or only trying it once. Urine toxicology results of those who used cannabis could be either negative or positive for THC. Participants with low cannabis use reported using cannabis between two and ten times across their lifetime, used less than once monthly in the past year, and/or denied recent cannabis use yet tested positive for THC. Moderate use was defined as cannabis consumption occurring one to three times per month over the past year. Heavy use was defined as using cannabis weekly to daily and/or meeting DSM-5 criteria for cannabis use disorder (CUD).

**Supplementary Table S1.** Adolescent cannabis use quantification and classification

| Cannabis Use Severity | Estimated frequency from clinician-elicited and self-reported measures | Met DSM-5 criteria for CUD | Urine toxicology for THC |
|---|---|---|---|
| **Adolescents who Did Not Use Cannabis** | | | |
| Never Used | Never used | No | − |
| Tried Once | Once only, no repeat | No | − |
| **Adolescents who Used Cannabis** | | | |
| Low Use | 2-10 lifetime uses; or<br>Less than once a month in the past 12 months without specifying "tried once"; or<br>Denied on self-report but urine toxicology THC+ | No | ± |
| Moderate Use | Once to 3 times a month in the past 12 months | No | ± |
| Heavy Use | Once a week to daily use; and/or<br>Meet DSM-5 criteria for CUD | Yes for some | ± |

*Abbreviations:* CUD: Cannabis Use Disorder; DSM-5: Diagnostic and Statistical Manual of Mental Disorders, Fifth Edition; THC: $\Delta^9$-tetrahydrocannabinol.

*SM2. Dimensional Measures of Symptomatology*

**Supplementary Table S2.** Details of depression, anxiety, suicidality, and anhedonia scales

| Measure | Number of Items | Possible Answers per Item | Score Range |
|---|---|---|---|
| Children's Depression Rating Scale-Revised (CDRS-R) | 17 | Each statement is rated on a scale of 1 to 5 or 1 to 7.<br>Specific examples of severity are provided for each item. | 17 – 113<br>Higher scores indicate more severe depression. |
| Multidimensional Anxiety Scale for Children (MASC) | 39 | 0 = "never true about me"<br>1 = "rarely true about me"<br>2 = "sometimes true about me"<br>3 = "often true about me" | 0 – 117<br>Higher scores indicate worse anxiety. |
| Beck Scale for Suicide Ideation (BSSI) | 19<br>5 screening items<br>14 follow-up items | Each statement is rated on a 3-point scale of 0 to 2.<br>Any score above 0 in any of the first five screening items prompts the respondent to complete the additional 14 follow-up items. | 0 – 38<br>Higher scores indicate more severe suicidality. |
| Temporal Experience of Pleasure Scale (TEPS) | <u>TEPS-A (anticipatory pleasure)</u>: 10<br><u>TEPS-C (consummatory pleasure)</u>: 8 | 1 = "very false for me"<br>2 = "moderately false for me"<br>3 = "slightly false for me"<br>4 = "slightly true for me"<br>5 = "moderately true for me"<br>6 = "very true for me" | <u>TEPS-A:</u> 10 – 60<br><u>TEPS-C:</u> 8 – 48<br>Lower scores indicate lower levels of pleasure (higher levels of anhedonia). |

*SM3. Reward Flanker Task (RFT) Practice Session in Mock Scanner*

The practice RFT comprised four mock runs of unequal duration with gradually increasing difficulty. Participants were informed that no real money would be won in this session. To mimic the MRI environment, pre-recorded scanner acquisition noises were presented during mock runs 2, 3, and 4 while participants became accustomed to performing the task. Each participant's in-scanner response window was calibrated from their mean response time in mock run 4, multiplied by 1.5, and limited to a maximum of 1700 ms. **Supplementary Table S3** provides details of the practice session.

**Supplementary Table S3.** Mock-scanner practice sequence for the Reward Flanker Task

| Practice Run # | Objective | Trials (n) | Response window | Task progression notes | Scanner-noise playback |
|---|---|---|---|---|---|
| Run 1 | Familiarization with scanner setup, trial sequence, and button-presses | 10 | Unlimited (no deadline) | Practice only | No noise |
| Run 2 | Practice under shortened time limit | 5 | Shortened relative to mock Run 1 | Flanker display shortened | Ambient Cryogenic only |
| Run 3 | Abridged rehearsal of real task run | 5 | Deadline = 1700 ms (matches real task) | Stimulus timing and response duration as in real task | Ambient Cryogenic + EPI |
| Run 4 | Timed rehearsal of real task run with performance recording | 30 | Same as mock Run 3 | As in mock Run 3. Response time and accuracy recorded | Ambient Cryogenic + EPI |

**Notes:** EPI = echo-planar imaging (recorded scanner gradient noise). "Ambient cryogenic" denotes background cryo-pump noise playback.

*SM4. Effect Size Calculations of Significant fMRI Results*

Effect sizes of significant findings were computed from permutation-derived *t*-statistics and associated degrees of freedom obtained from the output of FSL PALM analyses. Per the recommendation of the PALM development team, for significant correlations (cannabis use frequency and depression severity), we reported Pearson's *r* computed from *t*-statistics and degrees of freedom using the following formula[2,3]: $r = \sqrt{\frac{t^2}{t^2+df}}$

*SM4.* Post Hoc *Sensitivity Analyses*

*Post hoc* sensitivity analyses were conducted using G*Power v3.1.9.7 software[4] for group comparisons (32 adolescents who used cannabis vs. 85 adolescents who did not use cannabis) and correlations across the full sample (N = 117) and within the group of adolescents who used cannabis (n = 32).

### Supplementary Results

#### *SR1. RFT Behavioral Analysis Results*

**Supplementary Table S4.** Descriptive results from behavioral analysis across the full sample

| | **0¢ cue** | **10¢ cue** | **50¢ cue** | **? cue** | **All trials** |
|---|---|---|---|---|---|
| Accuracy (%) | 86.2 ± 13.1 | 85.2 ± 11.5 | 87.0 ± 11.9 | 86.3 ± 10.3 | 86.2 ± 10.0 |
| Reaction Time (ms) | 702.1 ± 127.7 | 695.0 ± 132.5 | 691.2 ± 130.2 | 706.9 ± 128.6 | 701.7 ± 126.5 |

**Notes**: Reported as Mean ± Standard Deviation.

*SR2. Exploratory Sex-Stratified Results*

**Supplementary Figure 1.** Significant findings across sexes.

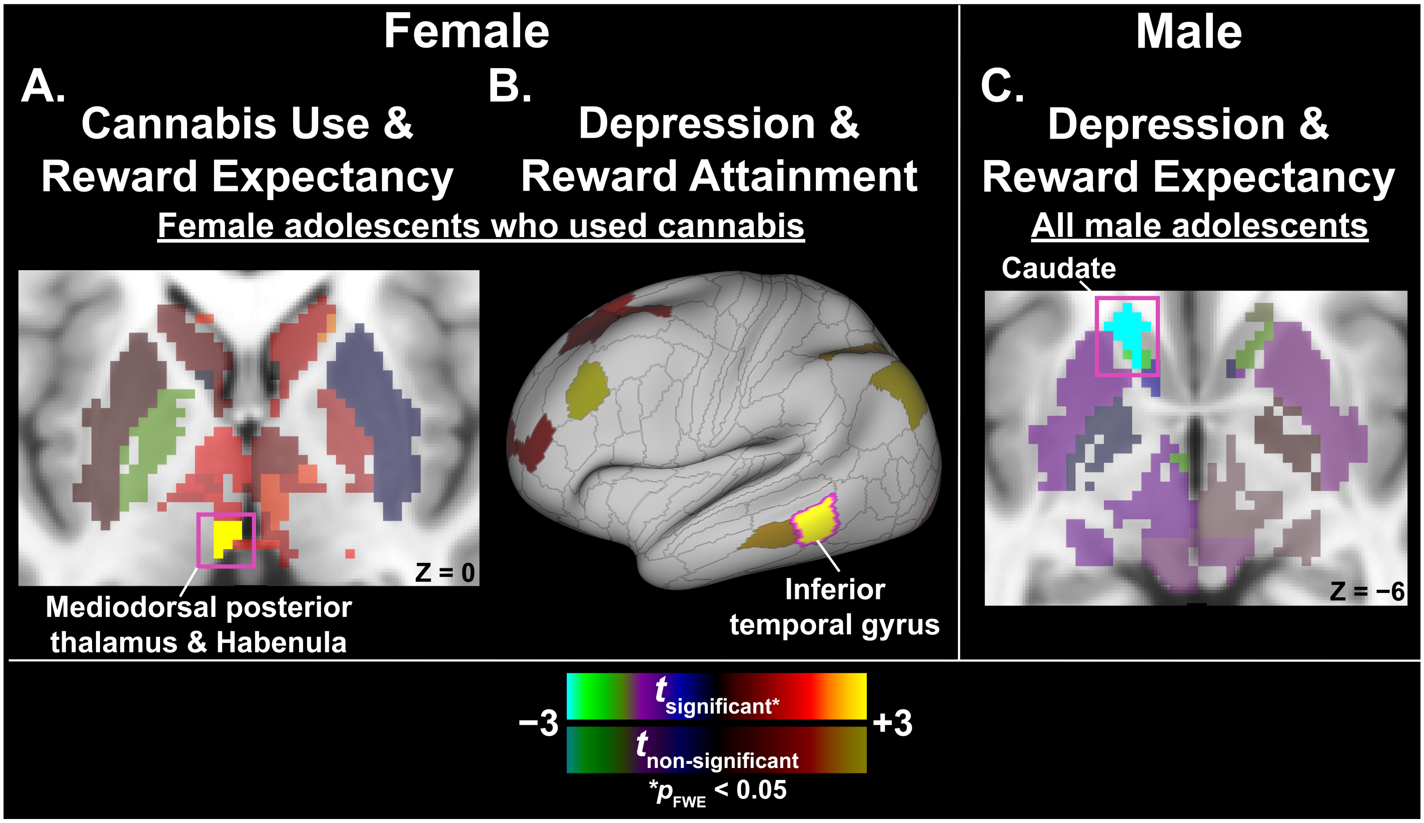


*Abbreviation:* FWE = family-wise error.

In female adolescents who used cannabis (n = 23), significant associations were observed between **(A)** cannabis use frequency and neural responses during reward expectancy, and **(B)** depression severity and neural responses during reward attainment. Across all male adolescents (n = 44), significant associations were detected between **(C)** depression severity and neural responses during reward expectancy. Results are displayed on axial slices through subcortical MNI space (**A** and **C**) and on the "very inflated" left cortical surface (**B**) in Connectome Workbench v2.0.0. Significant parcels (two-tailed $p_{FWE} < 0.05$) are outlined in fuchsia and labeled. Non-significant *t*-statistics are shown at 50% opacity relative to significant *t*-statistics.

**Supplementary Table S5.** Sex-stratified results

| RFT Contrast | Region (Lateralization) | HCP Cortical Label [a] | CAB-NP Label [b] | *t*-statistic [c] | Pearson's r [d] | $p_{FWE}$ |
|---|---|---|---|---|---|---|
| ***Female Adolescents with Cannabis Use** (n = 23) – **Cannabis Use Frequency*** [e] | | | | | | |
| *Reward Expectancy* | Mediodorsal posterior thalamus/habenula (L) | — | Visual-58 | 4.28 | 0.742 | 0.0159 |
| ***Female Adolescents with Cannabis Use** (n = 23) – **Depression Severity*** [f] | | | | | | |
| *Reward Attainment* | Inferior temporal gyrus (L) | TE1p | Frontoparietal_44 | 4.38 | 0.718 | 0.0282 |
| ***All Male Adolescents** (n = 44) – **Depression Severity*** [f] | | | | | | |
| *Reward Expectancy* | Caudate (L) | — | Visual-13 | −4.11 | −0.550 | 0.0144 |
| | Caudate (L) | — | Cingulo-Opercular-8 | −3.74 | −0.514 | 0.0381 |

*Abbreviations*: CAB-NP = Cole-Anticevic Brain-wide Network Partition; FWE = Family-wise error; HCP = Human Connectome Project; L = Left; R = Right; RFT = Reward Flanker Task.

[a] Labels per the HCP Cortical Atlas, as detailed in Glasser et al. (2016)

[b] Labels per Cole-Anticevic Brain-wide Network Partition v1.0.5 (equivalent labels per HCP S1200 Release cortical parcellation), which comprised 392 cortical nodes derived from HCP parcellation and 358 subcortical nodes from Cole-Anticevic atlas.

[c] Adjusted for age and head coil type.

[d] Pearson's *r* represents effect size and was computed from *t*-statistic and its corresponding degree of freedom output from FSL PALM.

[e] Cannabis use effect was adjusted for depression severity.

[f] Depression effect was adjusted for cannabis use frequency.

### Supplementary References

1. Nguyen TNB, Ely BA, Vitale A, et al. Cannabis Use is Related to Anhedonia in Adolescents With Diverse Mood and Anxiety Symptoms. *JAACAP Open*. 2025;doi:10.1016/j.jaacop.2025.02.003
2. Cohen J. *Statistical Power Analysis for the Behavioral Sciences*. 2013.
3. Rosenthal R, Rosnow RL. *Essentials of behavioral research: Methods and data analysis*. McGraw-Hill; 2008.
4. Erdfelder E, Faul F, Buchner A. GPOWER: A general power analysis program. *Behavior research methods, instruments, & computers*. 1996;28(1):1-11.
